# Mental Stress-Induced Peripheral Microvascular Dysfunction and Cardiovascular Events after a Premature Myocardial Infarction: A Prospective Cohort Study

**DOI:** 10.64898/2026.11.13.26368018

**Authors:** Viola Vaccarino, Hua She, Lisa Elon, Tené T. Lewis, Yi-An Ko, Nancy Murrah, Lucy H. Shallenberger, Lewam Stefanos, Tatum Roberts, Eric Fan-Lou, Bradley Pearce, Zachary T. Martin, Jose Medina Inojosa, Puja K. Mehta, Samaah Sullivan, J. Douglas Bremner, Paolo Raggi, Arshed Quyyumi, Amit J. Shah

**Affiliations:** Department of Epidemiology, Rollins School of Public Health, Emory University, Atlanta, GA, United States; Department of Medicine, Division of Cardiology, Emory University School of Medicine, Atlanta, GA, United States; Department of Biostatistics and Bioinformatics, Rollins School of Public Health, Emory University, Atlanta, GA, United States; Department of Exercise Physiology, College of Health Sciences and Professions, Ohio University, Athens, OH, United States; Diabetes Institute, Ohio University, Athens, OH, United States; The Institute to Advance Population Health (ADVANCE), Ohio University, Athens, OH, United States; Department of Epidemiology, Peter O’Donnell Jr. School of Public Health, University of Texas Southwestern Medical Center, Dallas, TX, United States; Department of Psychiatry and Behavioral Sciences, Emory University School of Medicine, Atlanta, GA, United States; Department of Radiology, Emory University School of Medicine, Atlanta, GA; Veterans Administration Medical Center, Decatur, Georgia, United States; Department of Medicine, Division of Cardiology, University of Alberta, Edmonton, Alberta, Canada

## Abstract

**Background:** Impaired macrovascular function during acute mental stress predicts adverse events in coronary artery disease, but microvascular stress responses are less understood. Because psychosocial stress and coronary microvascular dysfunction are more common in women, sex differences also warrant study.

**Methods:** We studied individuals aged 18-60 years within 8 months of a myocardial infarction (MI) who underwent a standardized mental stress challenge and were followed for cardiovascular events. We measured peripheral microvascular function with the reactive hyperemia index (RHI) before and after stress; abnormal RHI was defined as _≤_1.67. We also measured brachial flow-mediated vasodilation (FMD), interleukin-6 (IL-6), and myocardial ischemia. The primary endpoint was a composite of cardiovascular death, nonfatal MI, stroke, or heart failure hospitalization; a secondary endpoint included recurrent events. Cox regression examined stress-induced RHI decline, adjusting for pre-stress RHI and demographic, clinical, and psychological factors.

**Results:** Among 487 participants (mean age 51 years; 43% women), 89 first and 90 recurrent events occurred over a median 34 months. Women had lower RHI than men before and after stress, and a larger stress-induced decline. Each standard deviation stress-induced RHI decline was associated with 36% higher risk of first events (hazard ratio [HR], 1.36, 95% confidence interval [CI], 1.01, 1.82) and 44% higher risk of cumulative events (HR, 1.44, 95% CI, 1.04, 2.00), with no significant sex interaction. Abnormal post-stress RHI conferred adjusted HRs of 2.59 (95% CI, 1.13, 5.92) in women and 2.09 (95% CI, 1.11, 3.93) in men; abnormal pre-stress RHI was not predictive (HR, 1.12, 95% CI, 0.71, 1.76). IL-6 and FMD changes and stress-induced ischemia were related to RHI decline but did not explain its association with outcome.

**Conclusions:** Microvascular dysfunction with mental stress is more prevalent in women, and signals an elevated cardiovascular risk after MI in both women and men.

**What is already known on this topic:** Macrovascular dysfunction in response to acute mental stress has been related to increased risk of recurrent events in people with coronary artery disease. However, less is known about microvascular responses to stress.

**What this study adds:** A decline in microvascular function with mental stress is an independent predictor of subsequent cardiovascular events after a myocardial infarction (MI), and a stronger predictor than pre-stress microvascular function alone. Women have worse resting microvascular function and a more adverse microvascular response to stress.

**How this study might affect research, practice or policy:** Microvascular dysfunction may represent an important pathway linking psychological stress to adverse cardiovascular outcomes in people with MI. The microvascular response to stress should be evaluated as a potentially modifiable marker of cardiovascular risk and intervention studies should determine whether targeting stress-related microvascular dysfunction can reduce cardiovascular risk in this population.

## INTRODUCTION

Decades of research have uncovered the contribution of psychological stress to cardiovascular physiology and the development and progression of cardiovascular disease.^1^ In individuals with coronary artery disease (CAD), dynamic autonomic and molecular alterations during stress may help define cardiovascular risk.^2^ Hemodynamic and vascular responses to laboratory stress, including changes in brachial artery flow-mediated vasodilation (FMD), predict long-term prognosis in CAD beyond pre-stress levels.^3^ ^4^ Less is known, however, about microvascular responses to stress, which may capture distinct pathophysiological pathways not reflected by macrovascular measures.

Peripheral arterial tonometry is a non-invasive method to evaluate peripheral microvascular function by measuring arterial pulsatile changes in the fingertip during post-occlusive reactive hyperemia.^5^ The reactive hyperemia index (RHI) obtained with this device provides an estimate of peripheral microvascular vasodilator capacity that has been shown to predict cardiovascular events in individuals referred for evaluation to rule out CAD, patients with heart failure and those with stable CAD.^5^ ^6^ In an earlier, smaller wave of the Myocardial Infarction and Mental Stress (MIMS) study of young and middle-aged survivors of myocardial infarction (MI), stress-induced microvascular dysfunction predicted adverse events in women but not men.^7^ With a limited number of outcome events, that analysis may have lacked sufficient statistical power to determine whether this association extends to men. Here, leveraging the full MIMS sample across both recruitment waves, approximately doubling the number of participants and outcome events, we sought to examine whether the association between stress-induced peripheral microvascular dysfunction and adverse events after an MI generalizes to both sexes. We also aimed to characterize the clinical and psychosocial correlates of stress-induced peripheral microvascular dysfunction, and to identify potential intervening factors that may explain its link to adverse outcomes.

## METHODS

### Study Design and Participants

The present analysis draws on data collected across two enrollment waves (Wave 2 and Wave 3) of the MIMS prospective cohort study.^8^ ^9^ Both waves employed consistent protocols that were established after an initial pilot phase (Wave 1). Wave 2 recruited 313 individuals from April 2012 through March 2017; Wave 3 recruited 306 individuals from February 2020 through December 2023. Core study procedures and laboratory methods remained unchanged across waves.

Participants were recruited from Emory University-affiliated hospitals in Atlanta and were 18 to 60 years of age with a hospital admission for Type 1 MI within the prior 8 months, confirmed by medical record review using standard criteria.^10^ Exclusions included serious comorbid medical or psychiatric conditions, pregnancy or breastfeeding, immunosuppressive or non-antidepressant psychotropic medications, unstable cardiac conditions within 7 days, weight >450 lbs., and inability to safely withhold anti-ischemic medications for 24 hours. All participants underwent mental stress testing with myocardial perfusion imaging by single-photon emission computed tomography (SPECT).^11^ The study received approval from the Emory University Institutional Review Board, and each participant provided written informed consent prior to enrollment.

### Medical History and Other Clinical Characteristics

Trained staff collected sociodemographic data, medical history, blood pressure, and body mass index (BMI); race was self-reported. Clinical and angiographic data for the index MI were abstracted from medical records; obstructive CAD was defined as _≥_70% stenosis, and CAD burden was quantified with the Gensini score.^12^ Past-month angina frequency was assessed with the Seattle Angina Questionnaire, angina frequency subscale,^13^ and lifetime major depression and posttraumatic stress disorder (PTSD) with the Structured Clinical Interview for DSM-IV.^14^ Current psychological distress symptoms were measured with the Beck Depression Inventory-II,^15^ the PTSD Checklist, Civilian version,^16^ and the 4-item Perceived Stress Scale.^17^

### Mental Stress Testing

After a 30-minute resting baseline period, participants were exposed to a standardized public-speaking task designed to induce psychological stress.^8^ ^11^ After 2 minutes of preparation, participants delivered a 3-minute speech about an emotionally evocative scenario before a video camera and an evaluative panel. Heart rate and blood pressure were recorded every 5 minutes at rest and every minute during the speech.

### Peripheral Microvascular Function

RHI was measured before and 30 minutes after mental stress with the Endo-PAT2000 device (Itamar Medical, Caesarea, Israel), which records finger pulse volume amplitude (PVA).^6^ The reproducibility of RHI has been demonstrated before.^18^ Reactive hyperemia was induced by 5-minute upper-arm cuff occlusion at 50 mm Hg above systolic pressure. An automated algorithm derived the RHI as the ratio of post-deflation to baseline PVA in the occluded arm, normalized to the contralateral arm. Lower RHI indicates worse function; following existing guidelines for clinical significance, a threshold of RHI _≤_1.67 was used to define abnormal microvascular function.^19^

### Brachial Artery Endothelial Function

Brachial artery FMD was measured by B-mode ultrasound concurrently with RHI.^20^ Arterial diameter was measured with automated edge-detection software before and after 5-minute cuff occlusion.^20^ FMD was defined as the percentage diameter change within 60 seconds of cuff release; stress-induced change was calculated as post-stress minus resting FMD.

### Inflammation

Venous blood was drawn at rest and 90 minutes after stress, and plasma interleukin-6 (IL-6) was measured with a MesoScale Discovery electrochemiluminescence assay. IL-6 is acutely reactive to psychological stress,^21^ particularly in younger women,^22^ and has been associated with adverse cardiovascular outcomes in this population.^23^ Assay precision was acceptable, with an inter-assay coefficient of variation of 5.8% and an intra-assay coefficient of variation of 3.3% for IL-6 midpoint standards.

### Autonomic Function

Holter electrocardiograms (GE Marquette SEER; GE Medical Systems, Waukesha, WI) were recorded throughout the protocol, and high-and low-frequency heart rate variability (HRV) was derived in 5-minute rolling windows.^24^ ^25^ Only segments with a signal quality index of at least 0.70 and no evidence of atrial fibrillation were retained for analysis.

### Myocardial Perfusion Imaging

Participants underwent two SPECT scans with Technetium-99m (^99m^Tc) sestamibi using established protocols,^26^ one at rest and one with mental stress, the latter with tracer injection occurring one minute after the speech began. A blinded nuclear cardiologist (PR) scored 17 myocardial segments from 0 (normal) to 4 (absent perfusion); mental stress-induced ischemia was defined as a summed difference score _≥_3.^27^

### Outcome Assessment

Participants were contacted annually through October 31, 2019 (Wave 2) and May 31, 2026 (Wave 3). Events were ascertained through participant contact, medical records, and death registries, and adjudicated by blinded cardiologists. The primary endpoint was major adverse cardiovascular events (MACE) including cardiovascular death, nonfatal MI (Fourth Universal Definition, Type 1), stroke, or heart failure hospitalization.^10^ Since consideration of recurrent events may give insight on the true burden of cardiovascular disease, a secondary endpoint included first and recurrent events.

### Statistical Analysis

The main predictor was stress-induced RHI decline. Because restricted cubic splines showed no departure from linearity **(Supplemental Figure 1**), it was analyzed continuously, standardized to a mean of 0 and a standard deviation (SD) of 1. Baseline characteristics and physiological stress responses were compared according to presence or absence of abnormal post-stress RHI (≤1.67).^19^

Cox proportional hazards models examined time to first MACE, examining post-stress RHI decline adjusted for pre-stress RHI and study wave; the proportional hazards assumption was satisfied based on Schoenfeld residuals. Sex-specific hazard ratios (HR) and 95% confidence intervals (CI) were derived by fitting an interaction term between post-stress RHI decline and sex. Sequential, a priori defined models, adjusted for pre-stress RHI (Model 1); then adding age, sex, and race (Model 2); then adding ever smoking, BMI, hypertension, diabetes, left ventricular ejection fraction, and ST-segment elevation MI (Model 3); and then adding depression and PTSD symptoms (Model 4). Sensitivity analyses added marital status, education, medications, lifetime depression and PTSD, and perceived stress.

To illustrate the results, we plotted cumulative incidence curves according to presence or absence of abnormal post-stress RHI defined as RHI _≤_1.67, in the overall population and by sex, adjusting for all variables in Model 4.

For the secondary endpoint of recurrent events over time, we used the Andersen-Gill model for recurrent events, a generalization of the Cox model,^28^ and followed a similar sequential modeling strategy with the 4 nested models as for incident events.

To explore mechanisms, separate models added stress-induced changes in FMD and log-transformed IL-6, and stress-induced ischemia, and assessed whether there was attenuation of the RHI estimate; formal mediation analysis was not undertaken given the absence of attenuation. Biomarker data were missing for 2%–17% of participants owing to technical issues, unrelated to RHI, sex, or events, so these analyses used subsamples with complete data. Main-model covariates had <5% missing data; therefore no imputation was performed. Analyses used SAS version 9.4 (SAS Inc., Cary, NC).

## RESULTS

### Study Sample and Baseline Characteristics

Of 619 MIMS participants, 487 (79%; 290 from Wave 2, 197 from Wave 3) had complete RHI data and formed the analytic sample. RHI data were missing more in Wave 3 because of technical issues due to changes in equipment; also more in women (26% vs. 18%) and participants >50 years (23% vs. 18%), but missingness was unrelated to CAD risk factors or MI severity. The sample included 209 women (43%); 59% self-identified as Black or African American, and mean age was 50.9 years (SD, 6.7). Mean time since MI was 4.5 months (SD, 1.9).

The mean pre-stress RHI was 1.87 (SD, 0.59), and the mean post-stress RHI was 1.85 (SD, 0.63). These metrics were approximately normally distributed. Pre-stress RHI was moderately correlated with post-stress RHI and post-stress RHI decline (Spearman r=0.54 and r=0.37, respectively, p<0.001). Overall, 195 (40%) participants had an abnormal RHI (_≤_ 1.67) at rest, and 219 (45%) had an abnormal RHI post stress. Of 219 participants with abnormal RHI post-stress, 89 (41%) did not have an abnormal RHI pre-stress.

Women had lower RHI than men at rest (1.74 vs 1.93, p=0.01) and post-stress (1.73 vs. 1.93, p<0.001), more often had abnormal RHI (**Table 1, Supplemental Table 1**), and had a modestly larger RHI decline with stress (beta: 0.13, 95% CI, 0.03, 0.23, p=0.01, ∼0.2 SD). Participants with abnormal post-stress RHI were more often Black and had lower income and education, more symptoms of depression, PTSD, and perceived stress, higher BMI, more hypertension and dyslipidemia, and more angina (**Table 1**). Lifetime prevalence of depression and PTSD, history of MI prior to index event, index MI severity indicators, and medication use were similar. In contrast to post-stress abnormal RHI, abnormal pre-stress RHI was not related to psychological distress, and associations with metabolic risk factors other than BMI were attenuated (**Supplemental Table 1**).

**Table 1.** Baseline correlates of abnormal vs. normal peripheral microvascular function post-stress.

|  | Normal Post-Stress RHI (>1.67)<br>N=268 | Abnormal Post-Stress RHI (≤1.67)<br>N=219 | P | No. Missing |
|---|---|---|---|---|
| **Sociodemographic Factors** |  |  |  |  |
| Sex (female), % | 37 | 51 | 0.002 | 0 |
| Age, years, mean (SD) | 51.2 (6.7) | 50.5 (6.6) | 0.24 | 0 |
| Black, % | 49 | 72 | <0.001 | 0 |
| Income < \$25,000, % | 25 | 38 | 0.002 | 31 |
| Education >12 years, % | 68 | 57 | 0.02 | 5 |
| Married or living with partner, % | 52 | 39 | 0.005 | 0 |
| **Psychological Factors** |  |  |  |  |
| Lifetime history of major depression, % | 35 | 35 | 0.84 | 8 |
| Lifetime history of PTSD, % | 15 | 20 | 0.18 | 9 |
| Beck Depression Inventory, mean (SD) | 10.3 (9.4) | 13.0 (10.8) | 0.006 | 7 |
| PTSD Checklist, mean (SD) | 29.5 (13.3) | 33.4 (14.9) | 0.003 | 15 |
| Perceived Stress Scale, mean (SD) | 5.4 (3.6) | 6.3 (3.9) | 0.009 | 10 |
| **Medical History and Cardiovascular Risk Factors** |  |  |  |  |
| BMI, kg/m<sup>2</sup>, mean (SD) | 30.2 (5.9) | 32.6 (7.5) | <0.001 | 0 |
| Ever smoker, % | 48 | 51 | 0.47 | 7 |
| History of hypertension, % | 72 | 87 | <0.001 | 0 |
| History of dyslipidemia, % | 75 | 83 | 0.04 | 0 |
| History of diabetes, % | 32 | 37 | 0.26 | 0 |
| Angina frequency in past month, on scale 0-100 (SD)* | 12.7 (17.3) | 18.8 (20.7) | 0.001 | 19 |
| History of MI prior to index MI, % | 20 | 21 | 0.76 | 3 |
| History of revascularization prior to, or at index MI, % | 88 | 81 | 0.05 | 0 |
| **Clinical Characteristics** |  |  |  |  |
| ST-segment elevation MI, % | 34 | 28 | 0.15 | 8 |
| LV ejection fraction (%), mean (SD) | 50.2 (11.8) | 50.3 (12.4) | 0.95 | 0 |
| Gensini CAD severity score, log mean (SD)† | 3.1 (1.4) | 3.1 (1.5) | 0.80 | 23 |
| Obstructive CAD (stenosis ≥70%), %† | 83 | 84 | 0.90 | 23 |
| Three-vessel disease (at ≥70%), %† | 12 | 10 | 0.54 | 23 |
| **Medications** |  |  |  |  |
| Beta Blocker, % | 87 | 84 | 0.39 | 3 |
| Statins, % | 89 | 85 | 0.14 | 3 |
| Aspirin, % | 86 | 81 | 0.12 | 3 |
| ACE Inhibitors, % | 37 | 43 | 0.16 | 4 |
| ARB medications, % | 20 | 19 | 0.93 | 3 |
| Antidepressants, % | 16 | 13 | 0.24 | 4 |

SD: standard deviation; BMI: Body mass index; ACE: angiotensin-converting enzyme; ARB: angiotensin receptor blocker; MI: myocardial infarction; LV: left ventricle; CAD: coronary artery disease; RHI: reactive hyperemia index.
\* The Angina Frequency Subscale of the Seattle Angina Questionnaire was reverse coded from 0 (no angina episodes) to 100 (worst possible angina status).
† Angiographic data available in 464 participants (95%).

Participants with abnormal post-stress RHI had higher resting blood pressure but similar blood pressure responses to stress (**Table 2**). They also had lower resting FMD, a larger FMD decline and IL-6 increase with stress, and more stress-induced ischemia, but similar HRV.

**Table 2.** Stress response correlates of abnormal peripheral microvascular function post-stress.

|  | Normal Post-Stress RHI<br>(>1.67)<br>N=268 | Abnormal Post-Stress RHI<br>(≤1.67)<br>N=219 | P | No.<br>Missing |
|---|---|---|---|---|
| **Hemodynamics** |  |  |  |  |
| Systolic blood pressure, mm Hg |  |  |  |  |
| Rest | 129 (127, 132) | 135 (132, 138) | 0.003 | 3 |
| Change with mental stress<sup>*</sup> | 42 (40, 44) | 41 (39, 43) | 0.62 | 6 |
| Diastolic blood pressure, mm Hg |  |  |  |  |
| Rest | 81 (79, 82) | 84 (82, 86) | 0.007 | 3 |
| Change with mental stress* | 28 (27, 29) | 29 (27, 30) | 0.55 | 6 |
| Heart rate, beats/min |  |  |  |  |
| Rest | 64 (63, 65) | 64 (63, 66) | 0.83 | 3 |
| Change with stress<sup>*</sup> | 25 (23, 26) | 25 (23, 27) | 0.99 | 7 |
| **Macrovascular Function** |  |  |  |  |
| FMD, rest | 3.57 (3.20, 3.93) | 2.76 (2.34, 3.18) | 0.004 | 37 |
| FMD, change with mental stress† | -0.65 (-1.00, -0.29) | -1.23 (-1.64, -0.82) | 0.03 | 39 |
| **Inflammation** |  |  |  |  |
| lnIL-6, rest, pg/mL | 0.52 (0.44, 0.60) | 0.53 (0.44, 0.62) | 0.85 | 42 |
| lnIL-6, change with mental stress, pg/mL‡ | 0.49 (0.43, 0.55) | 0.60 (0.53, 0.67) | 0.02 | 83 |
| **Autonomic Function** |  |  |  |  |
| High-frequency HRV, rest, ln(ms<sup>2</sup>) | 5.85 (5.70, 6.00) | 5.91 (5.74, 6.08) | 0.60 | 47 |
| High-frequency HRV, change with mental stress§, ln(ms<sup>2</sup>) | -0.44 (-0.53, -0.35) | -0.41 (-0.52, -0.31) | 0.72 | 51 |
| Low-frequency HRV, rest, ln(ms<sup>2</sup>) | 6.36 (6.22, 6.50) | 6.26 (6.09, 6.42) | 0.35 | 47 |
| Low-frequency HRV, change with mental stress§, ln(ms<sup>2</sup>) | -0.17 (-0.26, -0.08) | -0.21 (-0.32, -0.11) | 0.51 | 51 |
| **Myocardial Perfusion** |  |  |  |  |
| Mental stress-induced ischemia, % | 13 | 21 | 0.02 | 10 |
| Summed rest score | 3.33 (2.60, 4.07) | 3.63 (2.79, 4.48) | 0.59 | 9 |
| Summed difference score | 0.71 (0.48, 0.94) | 1.08 (0.81, 1.35) | 0.04 | 10 |

All data are means and 95% confidence intervals (CI), derived from models that included a fixed effect for study wave. All differences are adjusted for pretest values.
FMD, flow-mediated vasodilation; HRV, heart rate variability; RHI: reactive hyperemia index.
\* Difference between maximum value during stress and minimum value during rest, adjusted for pretest values.
† Difference between values at 30 minutes post stress and pre-stress value.
‡ Difference between values at 90 minutes post-stress and pre-stress values.
§ Difference between HRV during the 3-minute speech task, and HRV averaged during the 10 minutes before mental stress.

### Association of Stress-Induced Peripheral Microvascular Dysfunction with Adverse Cardiovascular Events

The median follow-up time was 34 months (interquartile range, 25–38 months), which was similar in women and men and in participants with and without abnormal post-stress RHI. Two participants could not be contacted at follow-up and therefore their follow-up time was censored shortly after the baseline visit.

There were 89 first MACE (33 nonfatal MIs, 14 strokes, 32 heart failure hospitalizations, and 10 cardiovascular deaths) and 90 recurrent events, for 179 total events (**Table 3**): 69 in women (40 first) and 110 in men (49 first).

**Table 3.** Number of first and recurrent cardiovascular events in the total population and by sex.

|  | **Total**<br>**N=487** | **Women**<br>**N= 209** | **Men**<br>**N = 278** |
|---|---|---|---|
| **Cardiovascular Death** | 10 | 4 | 6 |
| **Non-fatal MI** |  |  |  |
| Total number of first events | 33 | 15 | 18 |
| Total number of recurrent events | 15 | 5 | 10 |
| **Stroke** |  |  |  |
| Total number of first events | 14 | 8 | 6 |
| Total number of recurrent events | 9 | 2 | 7 |
| **Hospitalizations for Heart Failure** |  |  |  |
| Total number of first events | 32 | 13 | 19 |
| Total number of recurrent events | 66 | 22 | 44 |
| **Cardiovascular Death, MI, Stroke or Heart Failure** |  |  |  |
| Total number of first events | 89 | 40 | 49 |
| Total number of recurrent events | 90 | 29 | 61 |
| **Total Number of Events (First and Recurrent)** | 179 | 69 | 110 |

MACE risk increased approximately linearly with RHI decline (**Supplemental Figure 1**). Each SD larger RHI decline (∼0.6 RHI units) was associated with a 58% higher risk of first MACE (HR, 1.58, 95% CI, 1.22, 2.05) in Model 1 (**Table 4**). The model was adjusted for pre-stress RHI, which was also significant (HR, 0.76, 95% CI, 0.59, 0.98, per SD), with higher pre-stress values being protective. After adjustment for demographic and clinical factors, the association was slightly attenuated but significant (Model 3: HR, 1.38, 95% CI, 1.04, 1.84), whereas pre-stress RHI was no longer significant (HR, 0.88, 95% CI, 0.68, 1.14). Adjustment for depression and PTSD symptoms had little effect on the estimate (Model 4: HR, 1.36, 95% CI, 1.01, 1.82). Estimates were numerically larger in women, but sex interactions were not significant.

**Table 4.**
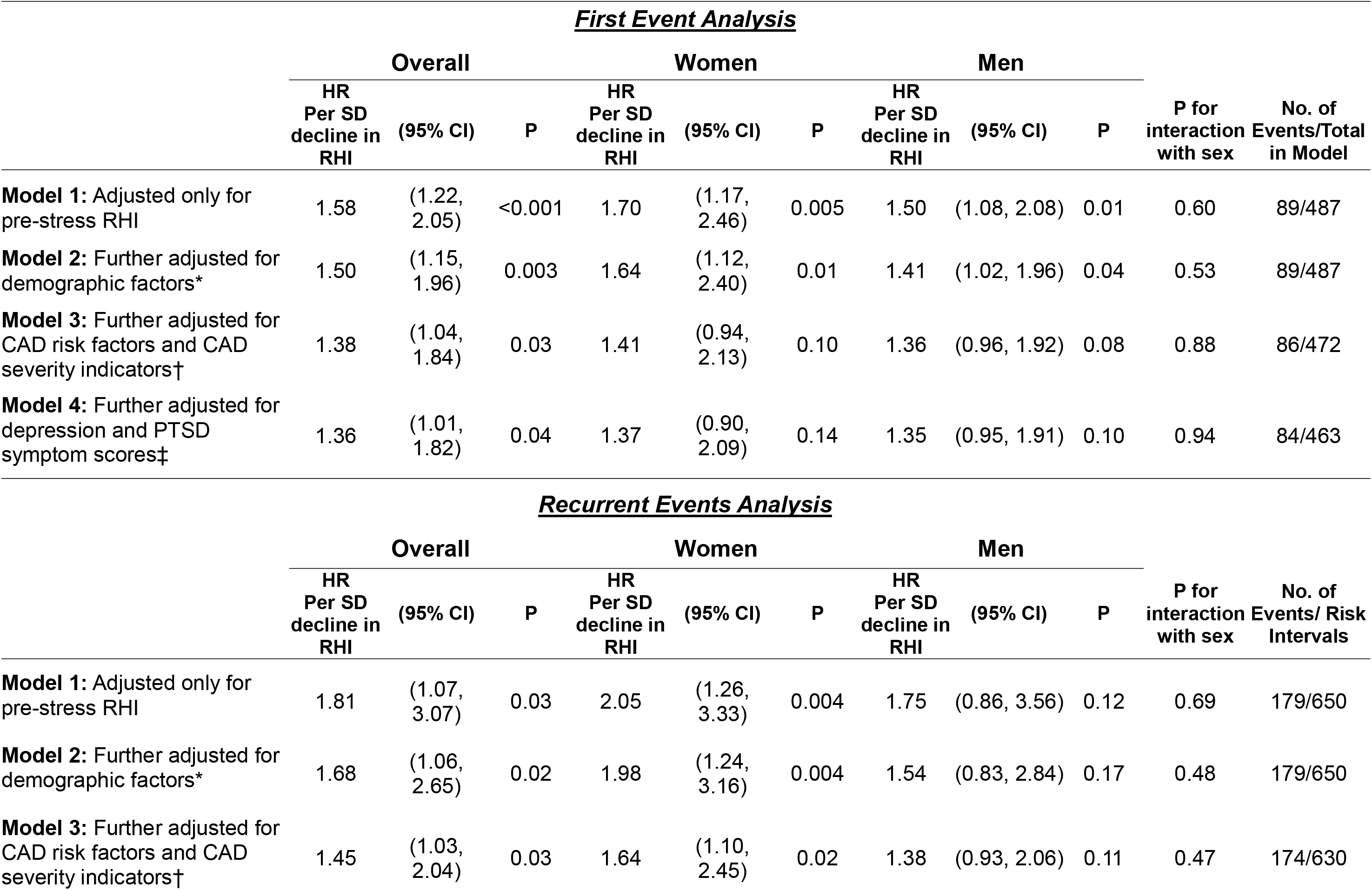
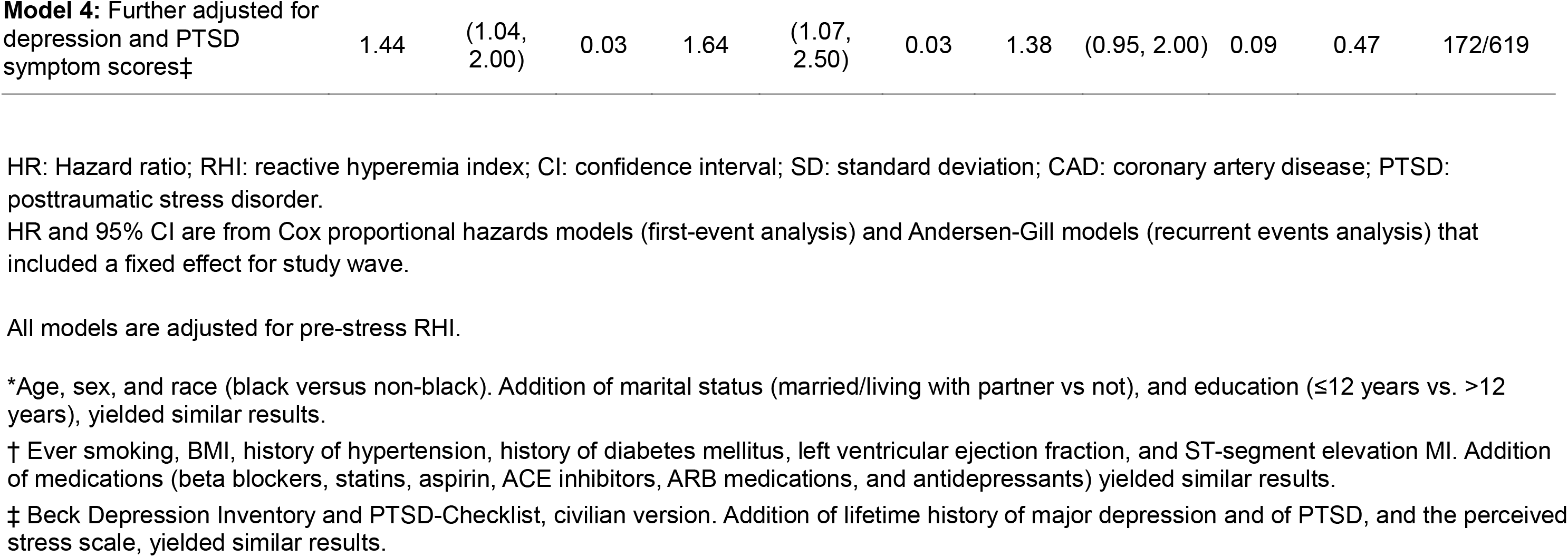
Association between stress-induced decline in peripheral microvascular function and the combined endpoint of cardiovascular death, nonfatal MI, stroke and hospitalizations for heart failure.

|  |  |  |  |  |  |  |  |  |  |  |  |
|---|---|---|---|---|---|---|---|---|---|---|---|
| ***First Event Analysis*** |  |  |  |  |  |  |  |  |  |  |  |
|  | **Overall** |  |  | **Women** |  |  | **Men** |  |  |  |  |
|  | **HR Per SD decline in RHI** | **(95% CI)** | **P** | **HR Per SD decline in RHI** | **(95% CI)** | **P** | **HR Per SD decline in RHI** | **(95% CI)** | **P** | **P for interaction with sex** | **No. of Events/Total in Model** |
| **Model 1:** Adjusted only for pre-stress RHI | 1.58 | (1.22, 2.05) | <0.001 | 1.70 | (1.17, 2.46) | 0.005 | 1.50 | (1.08, 2.08) | 0.01 | 0.60 | 89/487 |
| **Model 2:** Further adjusted for demographic factors\* | 1.50 | (1.15, 1.96) | 0.003 | 1.64 | (1.12, 2.40) | 0.01 | 1.41 | (1.02, 1.96) | 0.04 | 0.53 | 89/487 |
| **Model 3:** Further adjusted for CAD risk factors and CAD severity indicators† | 1.38 | (1.04, 1.84) | 0.03 | 1.41 | (0.94, 2.13) | 0.10 | 1.36 | (0.96, 1.92) | 0.08 | 0.88 | 86/472 |
| **Model 4:** Further adjusted for depression and PTSD symptom scores‡ | 1.36 | (1.01, 1.82) | 0.04 | 1.37 | (0.90, 2.09) | 0.14 | 1.35 | (0.95, 1.91) | 0.10 | 0.94 | 84/463 |
| ***Recurrent Events Analysis*** |  |  |  |  |  |  |  |  |  |  |  |
|  | **Overall** |  |  | **Women** |  |  | **Men** |  |  |  |  |
|  | **HR Per SD decline in RHI** | **(95% CI)** | **P** | **HR Per SD decline in RHI** | **(95% CI)** | **P** | **HR Per SD decline in RHI** | **(95% CI)** | **P** | **P for interaction with sex** | **No. of Events/ Risk Intervals** |
| **Model 1:** Adjusted only for pre-stress RHI | 1.81 | (1.07, 3.07) | 0.03 | 2.05 | (1.26, 3.33) | 0.004 | 1.75 | (0.86, 3.56) | 0.12 | 0.69 | 179/650 |
| **Model 2:** Further adjusted for demographic factors\* | 1.68 | (1.06, 2.65) | 0.02 | 1.98 | (1.24, 3.16) | 0.004 | 1.54 | (0.83, 2.84) | 0.17 | 0.48 | 179/650 |
| **Model 3:** Further adjusted for CAD risk factors and CAD severity indicators† | 1.45 | (1.03, 2.04) | 0.03 | 1.64 | (1.10, 2.45) | 0.02 | 1.38 | (0.93, 2.06) | 0.11 | 0.47 | 174/630 |
| **Model 4:** Further adjusted for depression and PTSD symptom scores‡ | 1.44 | (1.04, 2.00) | 0.03 | 1.64 | (1.07, 2.50) | 0.03 | 1.38 | (0.95, 2.00) | 0.09 | 0.47 | 172/619 |

HR: Hazard ratio; RHI: reactive hyperemia index; CI: confidence interval; SD: standard deviation; CAD: coronary artery disease; PTSD: posttraumatic stress disorder.
HR and 95% CI are from Cox proportional hazards models (first-event analysis) and Andersen-Gill models (recurrent events analysis) that included a fixed effect for study wave.
All models are adjusted for pre-stress RHI.
\*Age, sex, and race (black versus non-black). Addition of marital status (married/living with partner vs not), and education (≤12 years vs. >12 years), yielded similar results.
† Ever smoking, BMI, history of hypertension, history of diabetes mellitus, left ventricular ejection fraction, and ST-segment elevation MI. Addition of medications (beta blockers, statins, aspirin, ACE inhibitors, ARB medications, and antidepressants) yielded similar results.
‡ Beck Depression Inventory and PTSD-Checklist, civilian version. Addition of lifetime history of major depression and of PTSD, and the perceived stress scale, yielded similar results.

Recurrent event analyses yielded consistent, somewhat larger estimates (**Table 4**): each SD RHI decline was associated with 45% higher risk of first and recurrent events in Model 3 (HR, 1.45, 95% CI, 1.03, 2.04) and 44% higher risk of events in Model 4 (HR, 1.44, 95% CI, 1.04, 2.00). While estimates appeared stronger in women than men, the interaction with sex was not significant

In sensitivity analyses, results were similar after adding marital status and education to Model 2 (HR, 1.51, 95% CI, 1.15, 1.97), current medications to Model 3 (HR, 1.40, 95% CI, 1.04, 1.88), or lifetime depression, PTSD, and perceived stress to Model 4 (HR, 1.35, 95% CI, 1.01, 1.82).

When we examined post-stress RHI as a dichotomous variable, normal (RHI>1.67) vs. abnormal (RHI _≤_1.67), after adjusting for Model 4 variables and pre-stress RHI, the HR for MACE associated with abnormal RHI was 2.25 (95% CI, 1.32, 3.84) in the overall population (**Figure 1, Panel A**), 2.59 (95% CI, 1.13, 5.92) in women (**Figure 1, Panel B**), and 2.09 (95% CI, 1.11, 3.93) in men (**Figure 1, Panel C**). By contrast, an abnormal pre-stress RHI was not significantly related to outcome (HR, 1.12, 95% CI, 0.71, 1.76).

**Figure 1.**
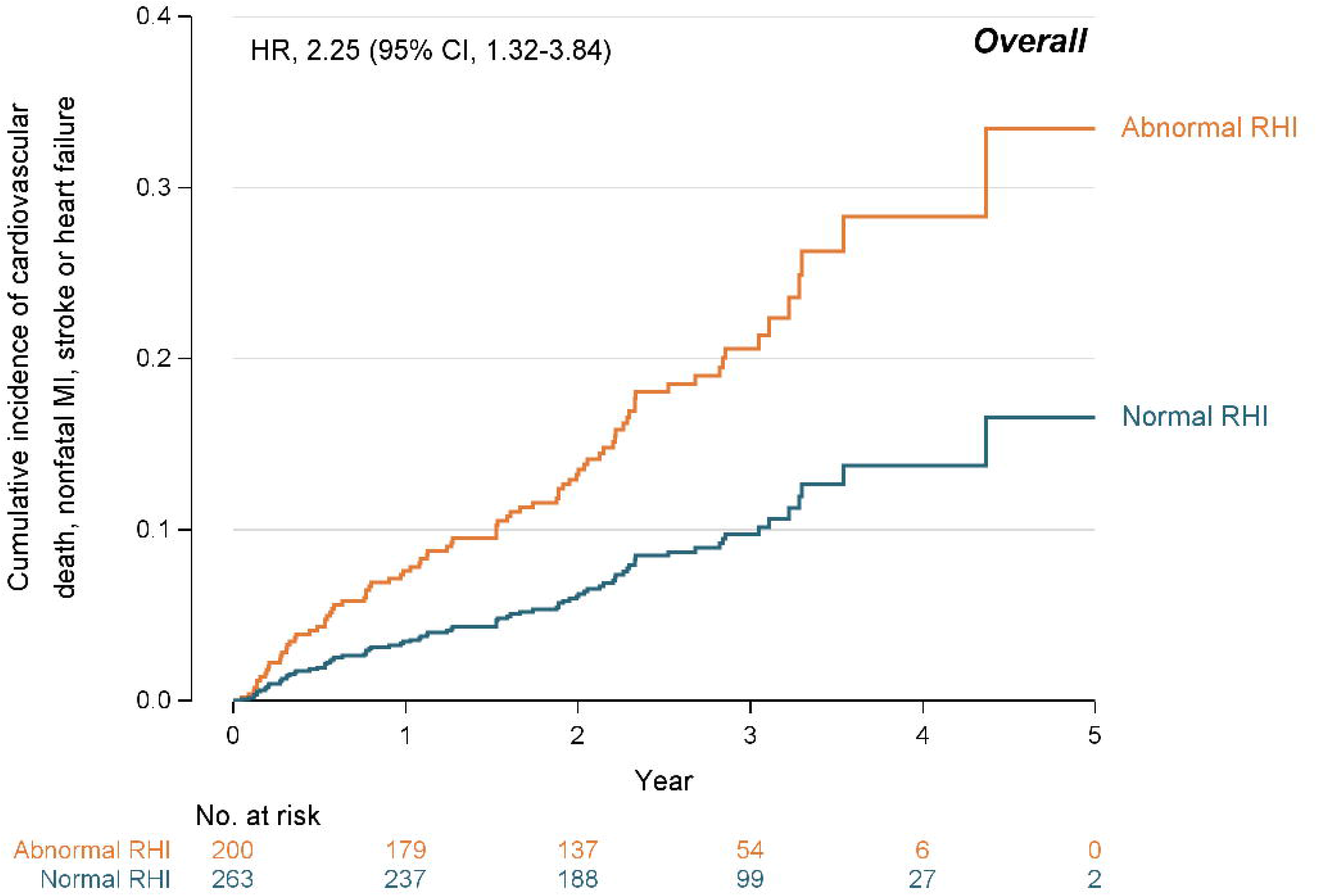
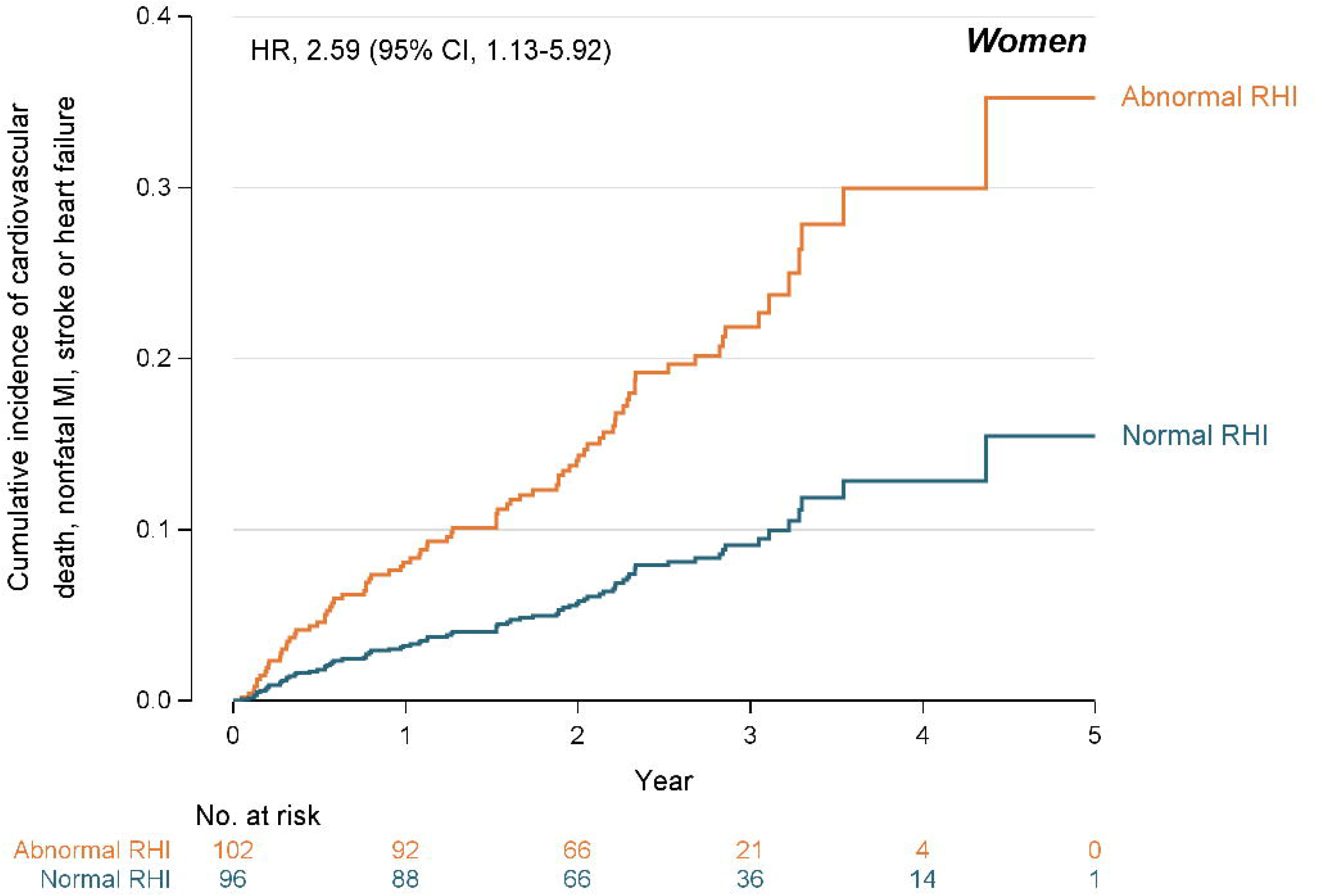
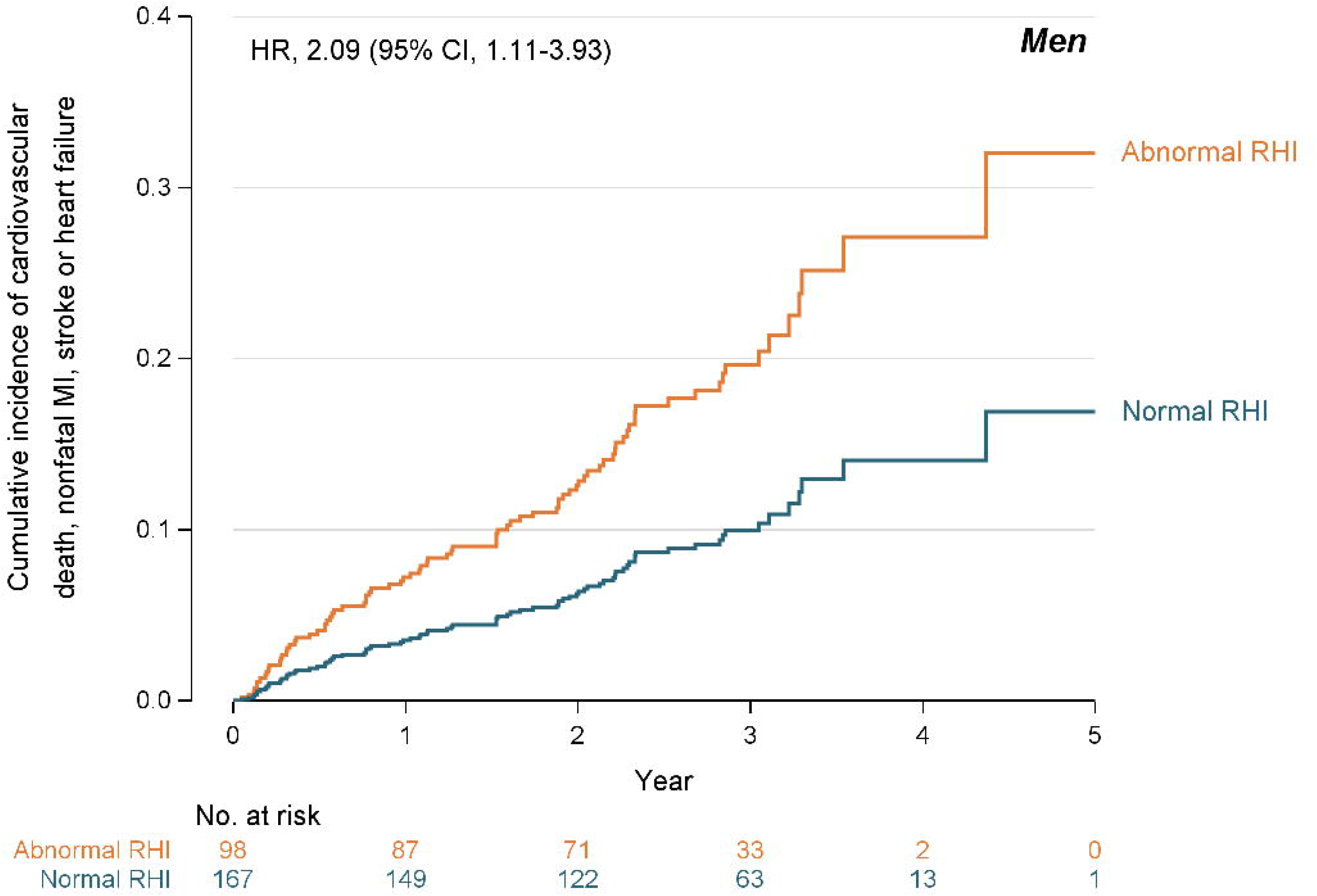
Adjusted Cumulative Incidence of the Primary Outcome (Cardiovascular Death, Nonfatal MI, Stroke or Hospitalizations for Heart Failure), by Abnormal (≤1.67) vs. Normal (>1.67) Reactive Hyperemia Index in the Overall Population and by Sex. Panel A: Overall Population; Panel B: Women; Panel C: Men. Graphs are derived from pooled Cox models that adjusted for Model 4 variables (age, sex (in overall graph), race, ever smoking, BMI, history of hypertension, history of diabetes mellitus, left ventricular ejection fraction, ST-segment elevation MI, Beck Depression Inventory score and PTSD-Checklist score).

FMD, IL-6, and myocardial perfusion data were available in 448 (92%), 404 (83%), and 477 (98%) participants, respectively, with unadjusted results similar to the main sample (**Supplemental Table 2**). Adjustment for stress-induced changes in FMD and IL-6 or for stress-induced ischemia did not materially change results, overall or by sex.

## DISCUSSION

Among individuals recently hospitalized for an MI, a decline in microvascular function induced by mental stress measured by reactive hyperemia was related to an increase in MACE risk in an approximately linear fashion. After adjusting for demographic, clinical and psychosocial factors, each SD decline of RHI post stress was associated with a 36% increased risk of incident MACE and 44% increased risk of incident and recurrent MACE over the follow-up. Although estimates in women appeared larger and more consistently significant than those of men, especially in recurrent event analysis, there was no significant interaction by sex, suggesting that the risk of MACE associated with stress-induced RHI decline is relevant to both women and men. An abnormal RHI post-stress, defined by a RHI value _≤_1.67, was associated with over two-fold increased MACE risk both in women and in men. By contrast, the association of pre-stress RHI with MACE was generally weaker and not statistically significant in this population. These results suggest that microvascular dysfunction may be a pathway linking psychological stress to adverse cardiovascular outcomes in populations with a significant burden of CAD, such as after a MI.

Prior work by our group and independent investigators has demonstrated that RHI is predictive of adverse cardiovascular events in individuals with CAD.^6^ ^29^ However up to now, much less has been known on whether dynamic changes of RHI with acute mental stress exacerbate risk in post-MI individuals above and beyond pre-stress levels. In an earlier publication based on the second wave of the MIMS study, microvascular dysfunction in response to mental stress showed an association with recurrent cardiovascular events in women but not in men.^7^ With approximately double the sample, we found that this risk extends to men. Women, however, had worse microvascular function before and after stress and a larger stress-induced decline, consistent with a high prevalence of microvascular dysfunction in women with suspected or established CAD than men, even without obstructive stenoses, and a more adverse microvascular response to stress.^30^

Despite comprehensive measurements encompassing behavioral, physiological and clinical factors, we were unable to elucidate the mechanisms underlying the excess risk associated with stress-induced microvascular dysfunction. An abnormal post-stress RHI was associated with a more adverse psychosocial risk profile, with a higher post-stress inflammatory response, with transient macrovascular dysfunction, and with a higher rate of mental stress induced ischemia. However, none of these measures explained the risk associated with a decline in RHI with stress, suggesting a role for unmeasured biological or psychosocial mechanisms that should be the object of future studies.

The robust association between post-stress RHI decline and MACE underscores the importance of capturing the psychological domain in post-MI patients and has important implications for future research and clinical care. The decline in RHI with laboratory mental stress may reflect susceptibility to stress-related microvascular dysregulation, which, if operative in daily life, could represent a mechanism linking chronic stress exposure to cardiovascular risk.^31^ Advances in mobile health technologies may soon enable the unobtrusive, continuous collection of extensive physiological data alongside real-world daily experiences, including ambulatory monitoring of hemodynamic, vascular, and autonomic function. Future large-scale prospective studies leveraging these technologies could help elucidate the mechanisms linking stress-induced microvascular dysfunction to subsequent cardiovascular events among patients recovering from an MI, and facilitate the validation and delivery of stress-reduction interventions. For example, there is emerging evidence that autonomic modulation using heart rate variability biofeedback can increase myocardial blood flow during mental stress compared with usual care.^32^ Although HRV was similar by RHI status in our study, this does not exclude sympathetic dysregulation, as HRV is an indirect measure of sympathetic activity.^25^

Strengths of this study include a racially diverse sample with many women, comprehensive assessments, excellent retention, and blinded event adjudication. Limitations include potential residual confounding and missing RHI data in 21% of participants, more often women, which may have led to underestimation of sex differences; however, missingness was unrelated to MI severity. Multiple tests were performed, although all were prespecified. We used a single stressor because sequential stressors may have cumulative effects, and a single stressor is necessary when using myocardial perfusion imaging with ^99m^Tc. Furthermore, whether the laboratory stressor used in our study generalizes to real-world stress exposures is an important constraint on interpretation. The single-center design with geographic concentration in Atlanta may limit the generalizability, but the diversity of the sample is a strength for generalizability relative to many post-MI cohorts.

In conclusion, microvascular dysfunction in response to mental stress signals an elevated risk of subsequent cardiovascular events in women and men after an MI, independent of measured sociodemographic, psychosocial and clinical characteristics. These findings underscore the importance of considering the microvascular response to stress as a potentially modifiable marker of cardiovascular risk in post-MI patients and highlight the need for intervention studies to determine whether targeting stress-related microvascular dysfunction can reduce this excess risk.

## Data Availability

All data produced in the present study are available upon reasonable request to the authors.

## Supplemental Material

Supplemental Table 1, Supplemental Table 2, Supplemental Figure 1.

