## Supplementary material for "Mental Stress-Induced Peripheral Microvascular Dysfunction and Cardiovascular Events after a Premature Myocardial Infarction: A Prospective Cohort Study": Suppl. Table 1, Suppl. Table 2, Suppl. Figure 1

**Supplemental Material**

**Supplemental Table 1.** Baseline correlates of abnormal vs. normal peripheral microvascular function pre-stress.

**Supplemental Table 2.** Association between mental stress induced decline in peripheral microvascular function and the combined endpoint of cardiovascular death, nonfatal MI, stroke and hospitalizations for heart failure after adding potentially intervening factors (macrovascular endothelial function, inflammation and stress-induced myocardial ischemia).

**Supplemental Figure 1.** Restricted-cubic-spline of the association between decline in peripheral microvascular function post-stress and cardiovascular outcomes (cardiovascular death, nonfatal MI, stroke or hospitalizations for heart failure).

| **Supplemental Table 1.**  Baseline correlates of abnormal vs. normal peripheral microvascular function pre-stress. | | | | |
| --- | --- | --- | --- | --- |
|  | **Normal Pre-Stress RHI (>1.67)**  **N=292** | **Abnormal Pre-Stress RHI (≤1.67)**  **N=195** | **P** | **No. Missing** |
| **Sociodemographic Factors** |  |  |  |  |
| Sex (female), % | 38 | 50 | 0.007 | 0 |
| Age, years, mean (SD) | 51.3 (6.4) | 50.4 (7.0) | 0.15 | 0 |
| Black, % | 50 | 72 | <0.001 | 0 |
| Income < $25,000, % | 27 | 36 | 0.049 | 31 |
| Education >12 years, % | 64 | 62 | 0.60 | 5 |
| Married or living with partner, % | 48 | 44 | 0.45 | 0 |
| **Psychological Factors** |  |  |  |  |
| Lifetime history of major depression, % | 33 | 38 | 0.22 | 8 |
| Lifetime history of PTSD, % | 16 | 19 | 0.45 | 9 |
| Beck Depression Inventory, mean (SD) | 10.8 (9.9) | 12.5 (10.5) | 0.10 | 7 |
| PTSD Checklist, mean (SD) | 30.2 (13.8) | 32.7 (14.5) | 0.08 | 15 |
| Perceived Stress Scale, mean (SD) | 5.5 (3.6) | 6.2 (3.8) | 0.06 | 10 |
| **Medical History and Cardiovascular Risk Factors** |  |  |  |  |
| BMI, kg/m^2^, mean (SD) | 30.5 (6.2) | 32.6 (7.4) | <0.001 | 0 |
| Ever smoker, % | 50 | 49 | 0.90 | 7 |
| History of hypertension, % | 77 | 82 | 0.23 | 0 |
| History of dyslipidemia, % | 76 | 82 | 0.17 | 0 |
| History of diabetes, % | 33 | 36 | 0.54 | 0 |
| Angina frequency in past month, on scale 0-100 (SD)* | 13.8 (18.8) | 17.7 (19.3) | 0.05 | 19 |
| History of MI prior to index MI, % | 20 | 20 | 0.87 | 3 |
| History of revascularization prior to, or at index MI, % | 88 | 80 | 0.02 | 0 |
| **Clinical Characteristics** |  |  |  |  |
| ST-segment elevation MI, % | 33 | 29 | 0.33 | 8 |
| LV ejection fraction (%), mean (SD) | 49.1 (12.4) | 52.0 (11.2) | 0.01 | 0 |
| Gensini CAD severity score, log mean (SD)† | 3.2 (1.4) | 3.0 (1.5) | 0.08 | 23 |
| Obstructive CAD (stenosis ≥70%), %† | 84 | 82 | 0.56 | 23 |
| Three-vessel disease (at ≥70%), %† | 11 | 13 | 0.44 | 23 |
| **Medications** |  |  |  |  |
| Beta Blocker, % | 87 | 84 | 0.37 | 3 |
| Statins, % | 91 | 81 | <0.001 | 3 |
| Aspirin, % | 86 | 81 | 0.10 | 3 |
| ACE Inhibitors, % | 39 | 40 | 0.90 | 4 |
| ARB medications, % | 18 | 22 | 0.38 | 3 |
| Antidepressants, % | 17 | 11 | 0.06 | 4 |

SD: standard deviation; BMI: Body mass index; ACE: angiotensin-converting enzyme; ARB: angiotensin receptor blocker; MI: myocardial infarction; LV: left ventricle; CAD: coronary artery disease; RHI: reactive hyperemia index.

* The Angina Frequency Scale from the Seattle Angina Questionnaire was reverse coded such that 0 indicates no angina episodes and 100 indicates the worst possible angina status.

† Angiographic data available in 464 participants (95%).

**Supplemental Table 2.** Association between mental stress induced decline in peripheral microvascular function and the combined endpoint of cardiovascular death, nonfatal MI, stroke and hospitalizations for heart failure after adding potentially intervening factors (macrovascular endothelial function, inflammation and stress-induced myocardial ischemia).

|  | **Overall** | | **Women** | | **Men** | |  |
| --- | --- | --- | --- | --- | --- | --- | --- |
|  | **HR Per SD Decline in RHI with Stress (95% CI)** | | | | | | No. of Events/Total in Model |
| **Participants with FMD data before and after stress** |  |  |  |  |  |  |  |
| Adjusted only for pre-stress RHI | 1.64 | (1.25, 2.16) | 1.75 | (1.18, 2.60) | 1.56 | (1.11, 2.20) | 81/448 |
| Further adjusted for FMD change with stress* | 1.63 | (1.24, 2.14) | 1.75 | (1.18, 2.58) | 1.55 | (1.10, 2.18) |  |
| **Participants with IL-6 data before and after stress** |  |  |  |  |  |  |  |
| Adjusted only for pre-stress RHI | 1.50 | (1.13, 1.99) | 1.80 | (1.17, 2.77) | 1.35 | (0.96, 1.90) | 67/404 |
| Further adjusted for IL-6 change with stress* | 1.52 | (1.14, 2.01) | 1.80 | (1.17, 2.77) | 1.37 | (0.97, 1.93) |  |
| **Participants with myocardial perfusion data before and after stress** |  |  |  |  |  |  |  |
| Adjusted only for pre-stress RHI | 1.57 | (1.20, 2.05) | 1.80 | (1.23, 2.64) | 1.42 | (1.02, 1.98) | 86/477 |
| Further adjusted for mental stress ischemia* | 1.56 | (1.19, 2.05) | 1.86 | (1.25, 2.77) | 1.38 | (0.99, 1.94) |  |

HR: Hazard ratio; CI: confidence interval; FMD: flow-mediated vasodilation; IL-6: interleukin-6; RHI: reactive hyperemia index.

All models are adjusted for pre-stress RHI.

* Results did not materially change if corresponding resting values were added to the models that included change values.

**Supplemental Figure 1.** Restricted-cubic-spline of the association between decline in peripheral microvascular function post-stress and cardiovascular outcomes (cardiovascular death, nonfatal MI, stroke or hospitalizations for heart failure).

-2

-1

0

1

2

Post-Stress RHI Decline

0

2

4

6

8

Hazard Ratio (95% CI)

Median

HR

95

% CI
